# Social-distancing Fatigue: Evidence from Real-time Crowd-sourced Traffic Data

**DOI:** 10.1101/2021.03.04.21252917

**Authors:** Jenni A. Shearston, Micaela E. Martinez, Yanelli Nunez, Markus Hilpert

**Affiliations:** Department of Environmental Health Sciences, Mailman School of Public Health, Columbia University, 722 West 168th St., New York, NY 10032, USA

**Keywords:** Traffic maps, human mobility, crowd-sourced data, social-distancing fatigue, coronavirus pandemic

## Abstract

**Introduction:** To mitigate the COVID-19 pandemic and prevent overwhelming the healthcare system, social-distancing policies such as school closure, stay-at-home orders, and indoor dining closure have been utilized worldwide. These policies function by reducing the rate of close contact within populations and results in decreased human mobility. Adherence to social distancing can substantially reduce disease spread. Thus, quantifying human mobility and social-distancing compliance, especially at high temporal resolution, can provide great insight into the impact of social distancing policies.

**Methods:** We used the movement of individuals around New York City (NYC), measured via traffic levels, as a proxy for human mobility and the impact of social-distancing policies (i.e., work from home policies, school closure, indoor dining closure etc.). By data mining Google traffic in real-time, and applying image processing, we derived high resolution time series of traffic in NYC. We used time series decomposition and generalized additive models to quantify changes in rush hour/non-rush hour, and weekday/weekend traffic, pre-pandemic and following the roll-out of multiple social distancing interventions.

**Results:** Mobility decreased sharply on March 14, 2020 following declaration of the pandemic. However, levels began rebounding by approximately April 13, almost 2 months before stay-at-home orders were lifted, indicating premature increase in mobility, which we term social-distancing fatigue. We also observed large impacts on diurnal traffic congestion, such that the pre-pandemic bi-modal weekday congestion representing morning and evening rush hour was dramatically altered. By September, traffic congestion rebounded to approximately 75% of pre-pandemic levels.

**Conclusion:** Using crowd-sourced traffic congestion data, we described changes in mobility in Manhattan, NYC, during the COVID-19 pandemic. These data can be used to inform human mobility changes during the current pandemic, in planning for responses to future pandemics, and in understanding the potential impact of large-scale traffic interventions such as congestion pricing policies.

**GRAPHICAL ABSTRACT:** 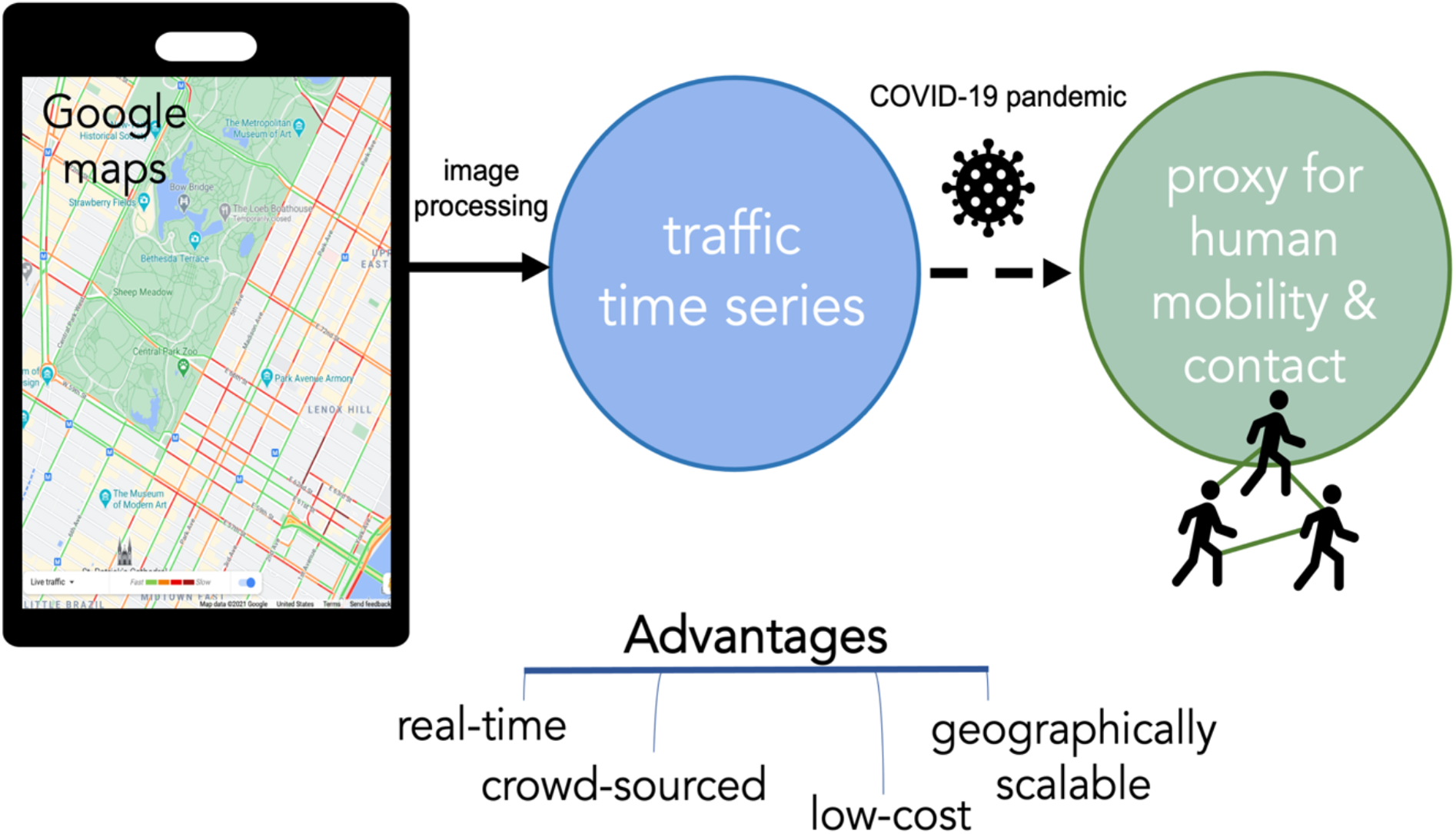

## 1. INTRODUCTION

The COVID-19 pandemic threatens almost every single country on Earth.^1,2^ To curb the pandemic and prevent overwhelming the healthcare system with COVID-19 cases, social-distancing policies such as school closures, non-essential business closures, curfews, and stay-at-home orders have been put into place. Social-distancing policies, specifically quarantines, are some of the oldest and most utilized strategies for epidemic control.^3^ Social distancing policies function by reducing physical contact and decreasing human mobility; adherence can substantially reduce transmission^4^ and case counts.^5^ Thus, quantifying human mobility and social-distancing compliance, especially at high temporal and spatial resolution, is critical for controlling the epidemic.

Several measures of social-distancing compliance have been used previously in the literature. Studies evaluating individual-level adherence to social-distancing policies frequently use self-report through online surveys.^6–8^ While these methods are very useful for evaluating determinants of adherence, they are not as useful for real-time epidemic monitoring because they are time-and labor-intensive and only capture a snapshot of the population’s behavior in a small sample at a single timepoint. In contrast, highly resolved data such as mobile phone call records or locations can be used to measure social-distancing^9^ and to inform mobility components of infectious disease transmission models.^10^ These measures, especially if publicly available, can contribute to epidemic monitoring. One such option is vehicular traffic condition smartphone apps, which base their maps on cellphone movement data. Traffic congestion maps are publicly available, updated in real-time, and can be used for public health research. For example, we have previously shown that colors in Google traffic maps correspond to relative vehicle speed, and used this information to infer traffic-related air pollution.^11^

App-based traffic information offers a pathway for assessing social-distancing, as increased traffic congestion is indicative of spending time outside the home that may result in opportunities for human interaction and contact. In fact, traffic data has been used to evaluate mobility during the pandemic in South Korea, finding that in some cities increases in traffic were correlated with increases in cases, but that in others traffic and cases were negatively correlated.^12^ Here we analyzed time series of traffic congestion that we derived from app-based maps. We quantify changes in traffic in the Manhattan borough of New York City (NYC) during the COVID-19 pandemic (January 1 to December 31, 2020). Our objective was to describe how social distancing interventions impacted population-level patterns of mobility throughout various stages of the pandemic.

## 2. METHODS

From January 1 through December 31, 2020, we obtained 12 tiles from Google traffic maps to view Manhattan’s entire street network every three hours in real-time. We used image processing methods, as described previously in Hilpert et al.,^11^ to identify the color-coded road segments. Color codes indicated four traffic categories: (i) green for free-flowing traffic, (ii) orange for medium traffic, (iii) red for traffic congestion, and (iv) maroon for severe traffic congestion. The colors are proxies for vehicle speed.^11,13^ In summary,^11^ map pixels for each of the four colors were counted and time series generated of the percent of the map area covered by each color.

To investigate changes in traffic congestion during the pandemic, we first characterized all four time series, but then restricted further analysis to the time series of red traffic congestion given its clear pandemic-related signal (e.g., decrease in April) and simpler interpretability (increases in red coverage indicate increases in congestion). We did this to avoid redundancy in analysis since all color coverage time series showed correlated temporal patterns. There were substantial changes in traffic congestion over the course of the pandemic and we captured this variation by partitioning the time series into four distinct time periods, which we refer to as COVID periods. We individually fit a generalized additive model (GAM)^14^ to each COVID period.

GAM models were constructed to predict the time series of percentage of total map area covered by traffic congestion, (*T*_*t*_) using two discrete predictor variables: hour of day, *h*_*t*_, which can assume values 0, 3, 6, 9, 12, 15, 18 and 21; and the binary variable *w*_*t*_ indicating whether a traffic map describes traffic on weekdays (*w*_1,*t*_) or during weekends (*w*_2,*t*_). We fitted the following model to the measured *T*_*t*_, *w*_1,*t*_, *w*_2,*t*_ and *h*_*t*_ data:

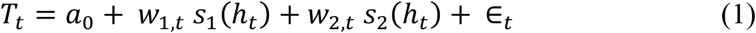

where *a*_0_ is the intercept (representing mean color coverage), and *s*_1_ and *s*_2_ are cyclic cubic regression splines with 8 degrees of freedom and zero mean. Degrees of freedom were selected using the generalized cross-validation criterion. The fitted models were then used to predict congestion levels hourly for a weekend and weekday, with 95% confidence intervals. Due to the morning and evening rush hour, traffic congestion displayed a 24-hour periodicity. The range of the diurnal fluctuation was measured as the deviation from the intercept.

To investigate potential social distancing fatigue (increased mobility before relaxation of stay-at-home policies), we applied seasonal decomposition of time series by Loess (STL)^15^ to the data from March 14 to June 16, a period including the *NY on PAUSE* policy intervention and the first stage of New York’s reopening (Phase 1). This analysis decomposed the time series into a periodic component, time trend, and the remainder (or residual). Prior to running the analysis, missing observations (n = 54) were imputed using predictions from the GAM models.

All analyses were conducted in R version 3.5.1 (R Foundation for Statistical Computing, Vienna, Austria). Simon Wood’s mgcv package was used to fit GAMs.^16^ STL analysis was completed using the ‘stl’ function in the stats package.

## 3. RESULTS

We found abrupt decreases in traffic congestion in Manhattan following school closures and the implementation of stay-at-home orders (*NY on PAUSE*). For a list of policies implemented by New York State and NYC in response to the COVID-19 pandemic, please see **Supplemental Table 1**. A map of the study area is shown in **Figure 1**, with colors representing the crowd-sourced traffic data. There was a dramatic decrease in traffic congestion after the implementation of social distancing policies. Stark differences in the proportion of green free-flowing traffic are readily apparent on the ring roads that trace the edges of Manhattan Island, as well as the main entry points, including the George Washington Bridge in the north and all tunnels and bridges in the south.

**Table 1.**
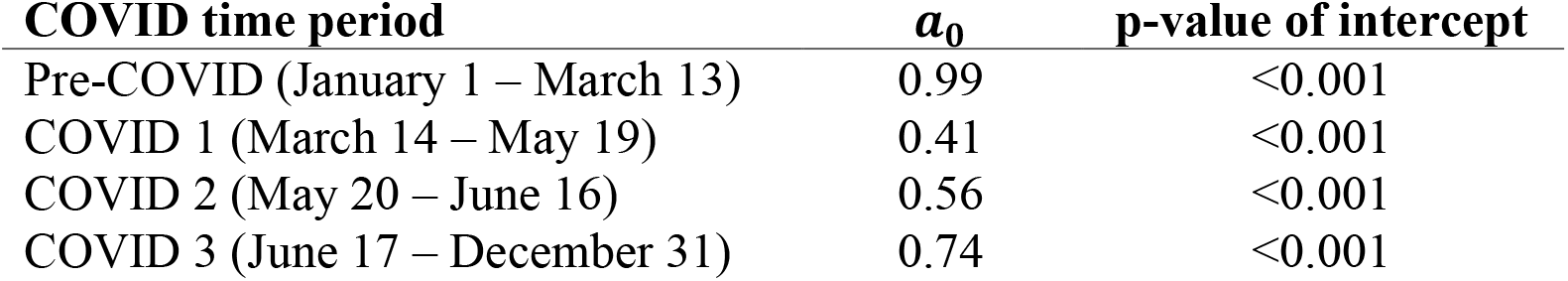
Intercepts *a*_0_ for generalized additive models describing the time series of red traffic congestion coverage *T*_*t*_ (% area) for four time periods during the COVID-19 pandemic

**Figure 1.**
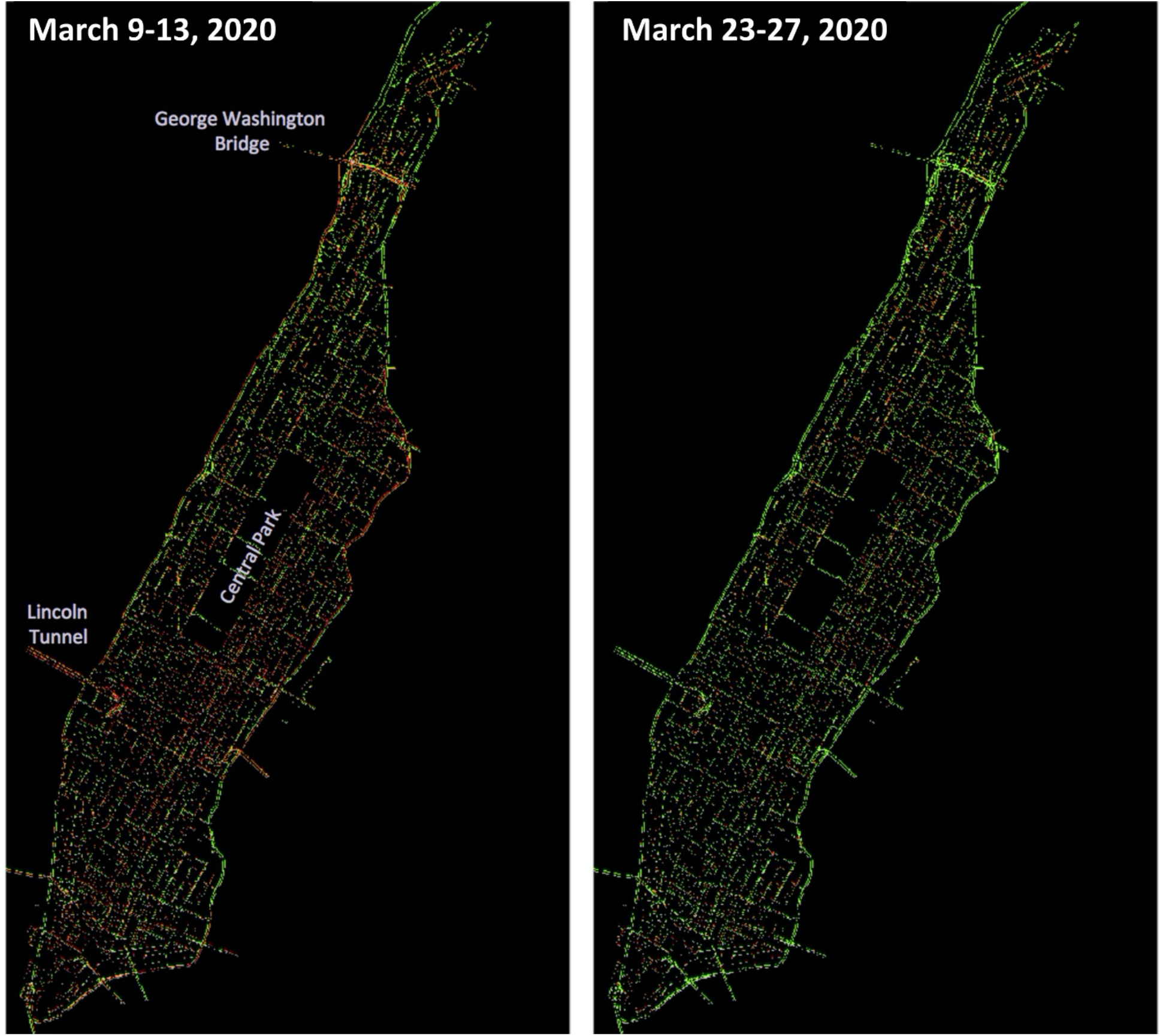
Pre- and Post-Pandemic Traffic. 5-day average traffic congestion in Manhattan during the 9:30 am weekday morning rush before (March 9-13) and after (March 23-27) school closures and stay-at-home orders (*NY on PAUSE* policies) went into effect in New York City. Red and orange indicates traffic congestion, while green indicates free-flowing traffic. After the implementation of *NY of PAUSE* policies, traffic became free flowing in the ring roads surrounding Manhattan, as well as the main entry points, including the George Washington Bridge in the north and all tunnels and bridges in the south.

Time series of the four congestion colors **(Figure 2)** reveal abrupt decreases in traffic congestion after March 14, the weekend before New York public school closure, with simultaneous increases in free-flowing traffic occurring. Importantly, we observed a steady increase in traffic congestion before New York on PAUSE ended on June 7th. This increase was most apparent in the red traffic congestion series, where traffic congestion appears to begin increasing on approximately May 1 and continues increasing up until about July 1. These increases occur before the end of stay-at-home orders and are indicative of social-distancing fatigue.

**Figure 2.**
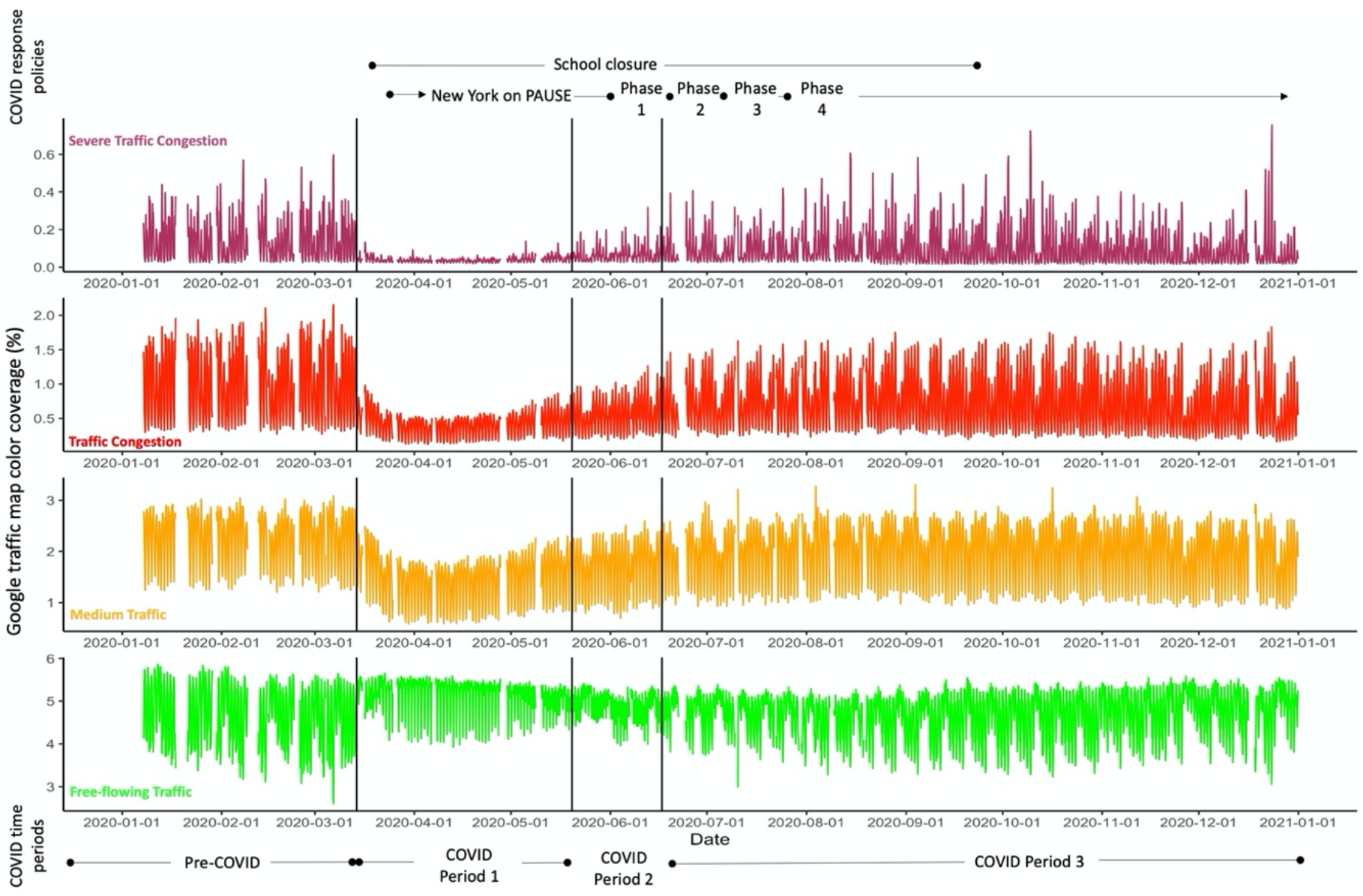
Time series of the percent of congestion color coverage within Manhattan for the four traffic categories. Vertical black lines delineate COVID time periods for the four separate GAM models, labeled below the bottom x-axis. The timeline of COVID response policies implemented by New York State are outlined at the top of the figure.

We identified four COVID Periods where traffic congestion changed markedly and fit each with a GAM model **(Equation 1)**. The Pre-COVID Period was defined as the start of the time series (January 1) through March 13, before congestion began to decrease dramatically. COVID Period 1 was defined as March 14 through May 19. Around May 19^th^ traffic congestion started to increase to the point that the model began poorly fitting the data; therefore, we defined COVID Period 2, which was May 20 through June 16, during which traffic congestion continued increasing. COVID Period 3 was June 17 through the end of the time series (December 31), where a more stable traffic pattern emerged.

Morning and evening rush hour impose a periodic cycle to traffic patterns, seen as daily peaks and troughs in traffic **(Figure 2)**. The pandemic not only caused changes in traffic trends, but also drove changes in the period cycle. In addition to the daily periodicity, there was also a weekly cycle where congestion was elevated during the five consecutive weekdays and was dampened during the two consecutive weekend days. The weekly cycle was most apparent in the Pre-COVID period and in the most recent period, COVID Period 3.

Mean percent area with red traffic congestion changed dramatically throughout the time period **(Table 1)**. Percent area with mean red traffic congestion (equivalent to the model intercept) was highest during the pre-COVID time period, and then decreased abruptly during COVID Period 1 (from a mean of 0.99% to 0.41%) before steadily increasing for COVID Periods 2 and 3. By COVID Period 3, the mean red traffic congestion area had rebounded to about 75% of the pre-pandemic average.

The rebound in red congestion coverage began far in advance of the implementation of Phase 1 reopening **(Figure 3, second panel)**. In STL analysis of COVID Periods 1 and 2, congestion appears to increase from approximately April 13^th^ onward, while Phase 1 reopening did not begin until June 8 (right dashed line).

**Figure 3.**
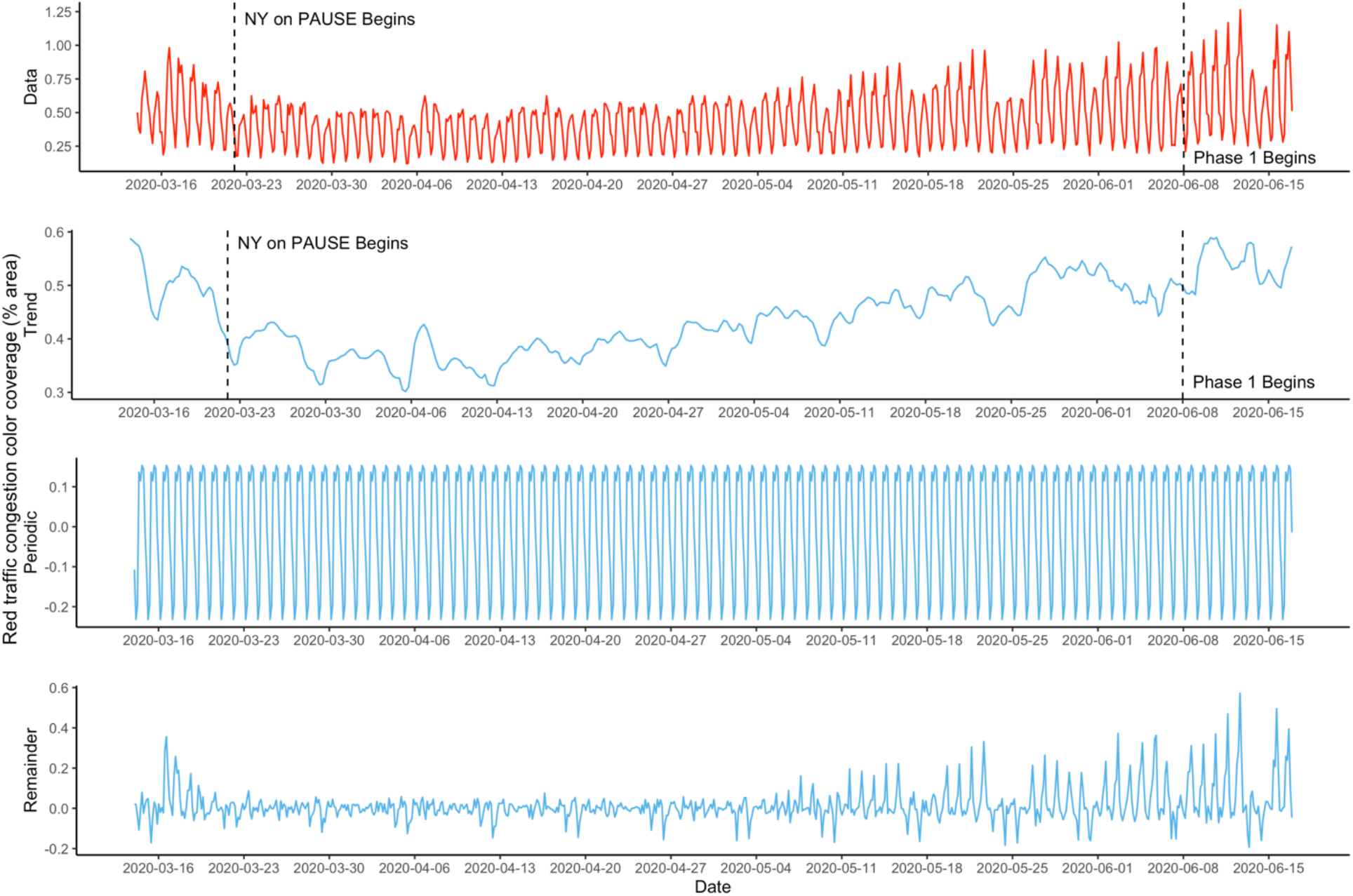
Time series decomposition of red traffic congestion from March 14 to June 16, 2020, with Loess. A time series of the red traffic congestion data is shown in the top panel, while the second panel shows the trend over time, the third panel shows the periodic daily pattern, and the bottom panel shows the remainder (or irregular) components of the time series. Dashed vertical lines indicate dates that policies were implemented to control the spread of the pandemic (left line: *NY on PAUSE*; right line: Phase 1 Reopening).

In addition to these changes in traffic trends, the pandemic substantially altered daily traffic periodicity, such that during the height of the pandemic in NYC in the Spring, there was little differentiation between weekday and weekend traffic patterns **(Figure 4, tan lines)**. During the Pre-COVID period (grey lines) rush hour peaks were highest, with weekdays demonstrating a clear bimodal distribution with peaks around 9am and 5pm, and weekends a clear unimodal peak around 5pm. However, during COVID Period 1, both weekday and weekend traffic peaks were greatly dampened, and the bimodal weekday distribution shifted to nearly unimodal, becoming very similar to the weekend distribution. During COVID Period 2 and 3 the daily traffic peaks were greater than for Period 1, but still lower than pre-pandemic levels. Even as overall traffic increased during these periods, the weekday distribution remained altered, such that the morning peak was much smaller than the evening peak.

**Figure 4.**
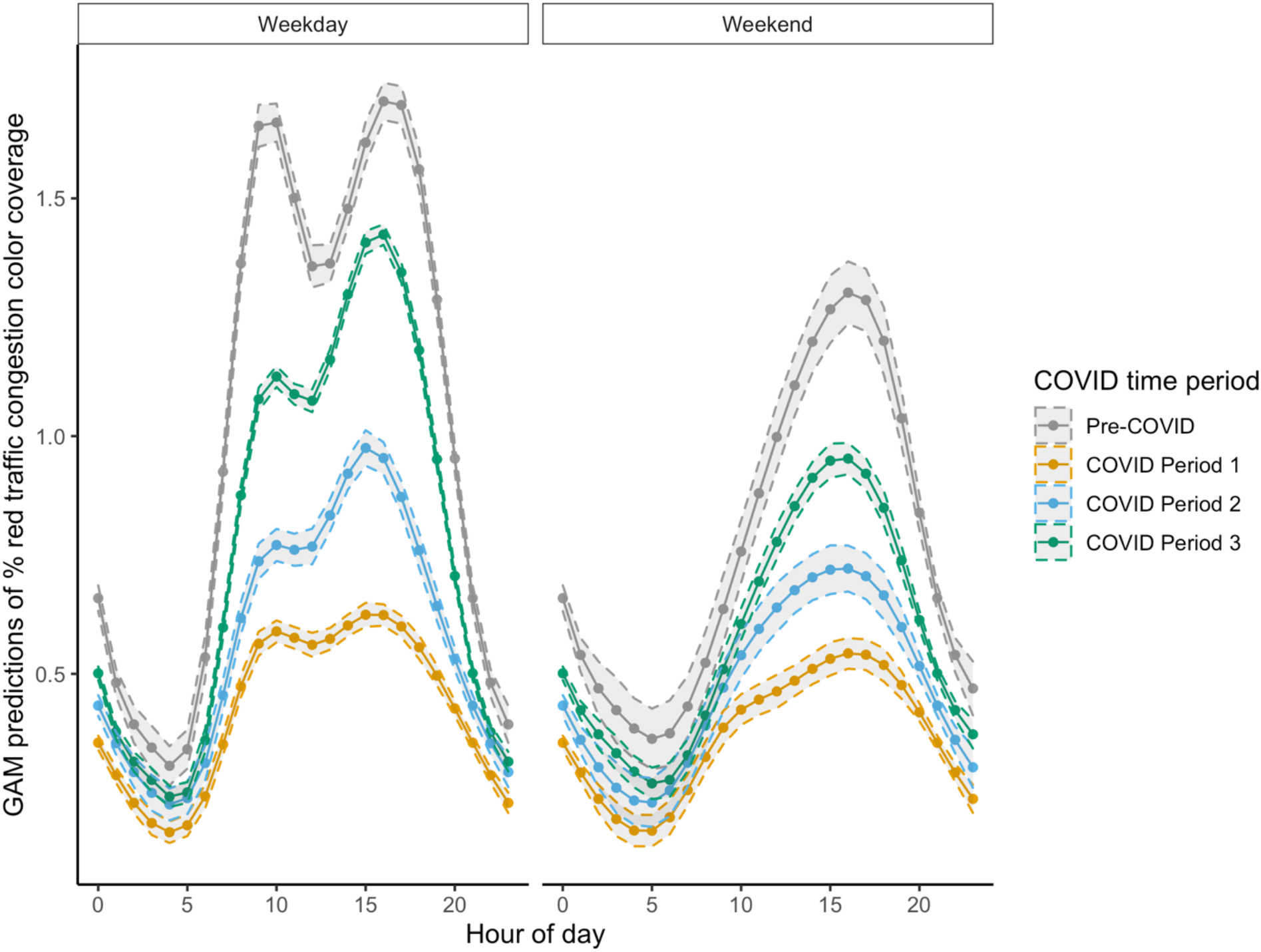
Diurnal fluctuation of red color coverage (in % area) on the traffic congestion maps for a typical weekday and weekend day (points and solid lines). Typical congestion levels are predicted from the GAM models with 95% confidence intervals (gray shading surrounding by dashed lines), for each COVID time period. Pre-COVID is from January 1 to March 13, COVID Period 1 is March 14 to May 19, COVID Period 2 is May 20 to June 16, and COVID Period 3 is June 17 to December 31.

A closer inspection of the GAM splines (**Supplemental Figure 1**), including the deviation of the fitted spline from the intercept, further highlight these differences in diurnal traffic congestion. In Panel C of Supplemental Figure 1, we can see that while the heights of the morning rush hour peak for weekdays remains highest for the Pre-COVID period, for COVID Period 3, the evening rush hour actually has a greater peak than the Pre-COVID period. This shift in traffic from the morning to the evening rush hour results in a diurnal congestion pattern that appears somewhere between the Pre-COVID weekday and weekend distributions.

## 4. DISCUSSION

We present evidence of social-distancing fatigue in Manhattan, NYC, from evaluation of traffic congestion levels during the COVID-19 pandemic. While traffic decreased sharply following the onset of the pandemic and implementation of response policies, levels were already rebounding almost two months before stay-at-home orders (*NY on PAUSE*) were lifted on June 8. Overall, we identified four COVID periods based on traffic congestion patterns. While the dates delineating the COVID period sometimes lined up with the implementation of official social distancing policies, they did not always. For example, traffic congestion began to plummet on March 14, but school closures were not implemented until March 16. NYC bars and restaurants closed in-person service on March 17, and *NY on PAUSE* began on March 22. The time period during which *NY on PAUSE* was in effect spans two different COVID Periods because traffic began to rebound far in advance of the first reopening policy (Phase 1 reopening) which occurred on June 8^th^. COVID Period 3, when traffic began to stabilize (June 17^th^), was very close to the Phase 2 re-opening date of June 22 (Figure 2). We also observe large impacts of the pandemic on the distribution of traffic throughout the day, such that the pre-pandemic bi-modal weekday diurnal congestion representing the morning and evening rush hour was dramatically depressed. By September, traffic congestion had rebounded to approximately 75% of pre-pandemic levels.

Dramatic decreases in traffic have been noted in NYC^17^ and many other places^18–21^ in response to the pandemic, including Padova, Italy^18^ and Auckland, New Zealand.^19^ The study in Padova found a decrease in traffic flow (vehicle counts per unit time) in 2020 of approximately 70%, compared to 2018-2017.^18^ Similarly, the study in Auckland found a 60-80% decrease in traffic flow following travel restrictions implemented in response to the pandemic.^19^ Another analysis of traffic changes in the NYC metro area found reductions of 66% in passenger vehicle traffic the week of March 28-April 3.^17^ While the traffic flows reported by these other studies cannot be directly compared to our metrics, our findings were in line with those previously published. We found a 59% reduction in total map area covered by traffic congestion during March 14-May 19, compared to January 1-March 13, and we were able to extent our analysis through the end of 2020.

In this paper, we use changes in traffic as a measure of human mobility and an indicator for social-distancing interventions. However, an increase in traffic congestion does not necessarily correlate with increased case counts of COVID-19, as a study evaluating correlations between traffic and cases in various cities of South Korea found.^12^ Broadly, we can conceptualize COVID-19 non-pharmaceutical interventions in four categories: face mask mandates, isolation or quarantine (including stay-at-home orders), traffic or travel restrictions (such as limiting travel between cities or counties), and social-distancing (closure of schools or other businesses, or limiting gatherings).^22^ A review found that social-distancing was the most effective single non-pharmaceutical intervention, while combining social-distancing with at least one other intervention was even more effective.^22^ Traffic congestion can be thought of as an indicator for three of these intervention groupings (all but mask mandates), as human mobility is a component of isolation or quarantine, traffic or travel restrictions, and social-distancing. As such, it makes sense to hypothesize that increased congestion may correlate with increased opportunity for SARS-CoV-2 transmission. However, increases in traffic congestion may also be an indicator of a switch in transportation methods in response to the pandemic; studies have documented dramatic decreases in use of public transportation in NYC^23^ and metro areas in Sweden.^24^ It is possible that individuals who otherwise would have used public transit such as subways or buses are now relying more heavily on private or shared vehicles to reduce their chance of exposure, as was found in Canada.^25^

Understanding changes in traffic congestion during the pandemic is useful not only as a proxy for human mobility, but also for implications on large-scale traffic interventions that may be implemented in the future. For example, in 2019 New York City became the first American city to approve a congestion pricing policy,^26^ with implementation scheduled to begin in early 2021 (although this was delayed). The large-scale traffic disruption that followed implementation of *NY on PAUSE*, however, may be useful in informing what kinds of changes in traffic patterns could reduce congestion. As we saw in COVID period 1, traffic congestion can be reduced both by depression of overall traffic and by dampening the bimodal diurnal congestion driven by weekday rush hour.

In this paper, use of STL analysis allowed for clear identification of the increasing trend in traffic congestion, while use of GAM models allowed us to account for the changing within-day distribution of the congestion data over the course of the pandemic. These methods allowed us to identify the presence of social-distancing fatigue and to describe traffic changes by hour, demonstrating a clearer picture of the impact of the pandemic on traffic in NYC. This information is useful not only as a proxy for tracking human mobility, but also for understanding how large-scale travel restrictions can impact traffic patterns and congestion overall.

Out study has some notable limitations. First, we include a fairly limited geographic area (the borough of Manhattan, NYC), and thus results of our study may not be generalizable to other areas. However, studies in other cities also find dramatic decreases in traffic, on a similar scale as that reported here.^18,19,21^ Second, our data source does not allow for disaggregation of passenger vehicles and trucks, which were likely differentially impacted by responses to the pandemic (as trucking operations may be considered essential businesses). Studies in NYC^27^ and Somerville MA^20^ have found decreases in both vehicles and trucks, though the decrease was substantially less for trucks. Third, we do not disaggregate traffic changes by neighborhoods, and thus do not report variation in congestion changes within Manhattan. We recommend that future studies describe and evaluate these potential differences.

## 5. CONCLUSIONS

Using highly temporally resolved, crowd-sourced traffic congestion data, we describe changes in traffic congestion in Manhattan, NYC, during the COVID-19 pandemic. We report dramatic declines in traffic congestion during the initial stages of the pandemic and implementation of stay-at-home orders, followed by rebounds in congestion nearly two months before stay-at-home orders were reversed, evidence of social-distancing fatigue. Additionally, we describe changes in diurnal traffic congestion patterns for weekdays and weekends, by hour, for four time periods during the pandemic. This data can be used to inform human mobility changes during the current pandemic, in planning for responses to future pandemics, as well as in understanding the potential impact of large-scale traffic interventions such as congestion pricing policies.

## Data Availability

Data is publicly available.

