## Supplementary material for "Social-distancing Fatigue: Evidence from Real-time Crowd-sourced Traffic Data": supp.table.1_supp.fig.1

**Supplemental Table 1.** Timeline of policies implemented by New York State (NYS) and City (NYC) in response to the COVID-19 pandemic

| Implementation Date | Policy |
|---|---|
| March 16, 2020 | NYC public schools close |
| March 17, 2020 | NYC bars and restaurants close, except for delivery |
| March 22, 2020 | NYS on Pause Program begins: all non-essential workers must stay home |
| June 8, 2020 | NYC begins Phase 1 of reopening |
| June 22, 2020 | NYC begins Phase 2 of reopening |
| June 24, 2020 | NY, NJ, and CT require travelers to self-quarantine for 14 days if traveling from hot spots |
| July 6, 2020 | NYC begins Phase 3 of reopening, without indoor dining |
| July 19, 2020 | NYC begins Phase 4 reopening, excluding malls, museums and indoor dining/bars |
| September 21, 2020 | NYC public schools open to modified in-person teaching |
| November 19, 2020 | NYC public schools re-close to in-person teaching |

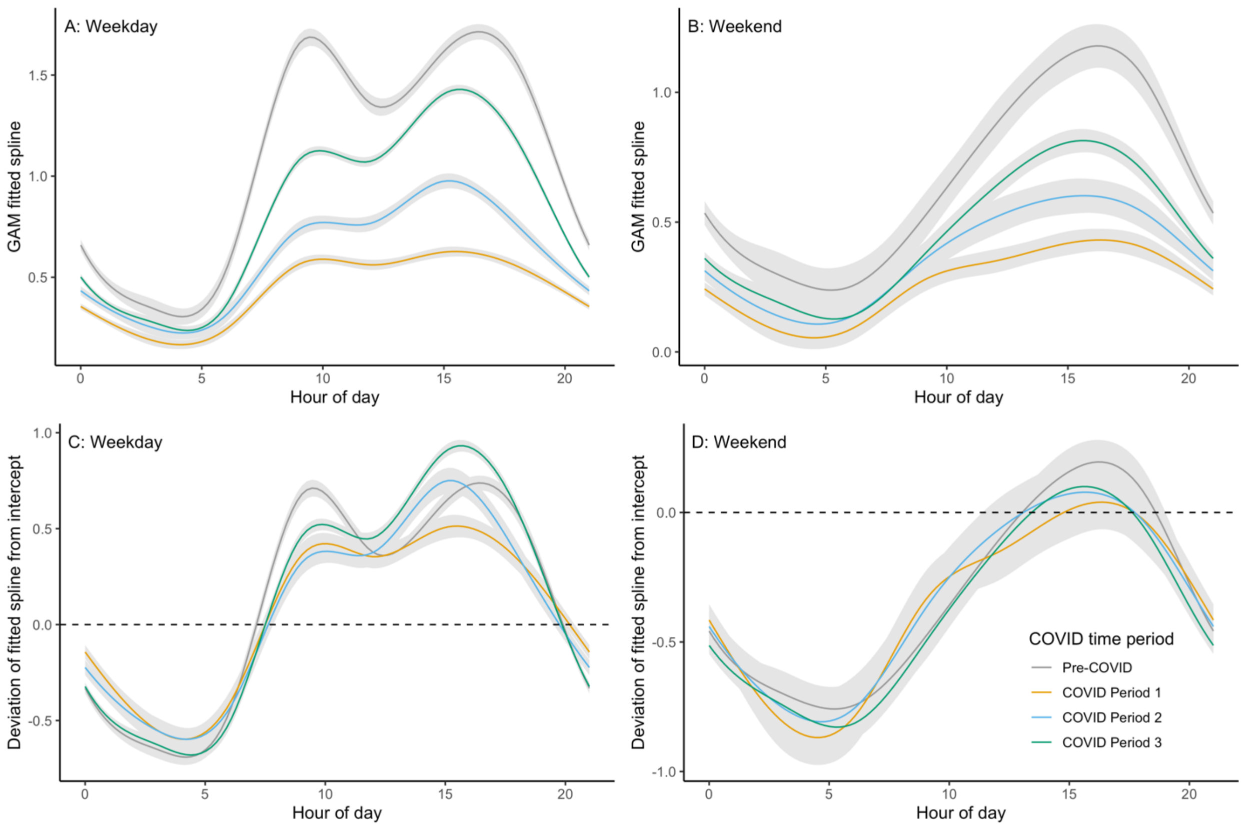


**Supplemental Figure 1.** Fitted splines from the GAM models for each COVID time period (each modelled time period is represented by a different color; gray shading indicates 95% confidence intervals). The top row shows GAM fitted splines for weekdays (Panel A) and weekends (Panel B). The bottom row shows the percent deviation of the fitted spline from the intercept for weekdays (Panel C) and weekends (Panel D). Pre-COVID is from January 1 to March 13, COVID Period 1 is March 14 to May 19, COVID Period 2 is May 20 to June 16, and COVID Period 3 is June 17 to December 31.
